# Epidemiological topology data analysis links severe COVID-19 to RAAS and hyperlipidemia associated metabolic syndrome conditions

**DOI:** 10.1101/2022.03.31.22273239

**Authors:** Daniel Platt, Aritra Bose, Chaya Levovitz, Kahn Rhrissorrakrai, Laxmi Parida

**Affiliations:** IBM Research, Yorktown Heights, NY

## Abstract

The emergence of COVID19 created incredible worldwide challenges but offers unique opportunities to understand the physiology of its risk factors and their interactions with complex disease conditions, such as metabolic syndrome. Epidemiological analysis powered by topological data analysis (TDA) is a novel approach to uncover these clinically relevant interactions. Here TDA utilized Explorys data to discover associations among severe COVID19 and metabolic syndrome, and it explored the probative value of drug prescriptions to capture the involvement of RAAS and hypertension with COVID19 as well as modification of risk factor impact by hyperlipidemia on severe COVID19.

## Introduction

Severe COVID-19 (C19) due to the SARS-COV-2 pandemic challenged medical services around the world. Metabolic syndrome represents a highly tangled contributor to severe C19 risk.^1^ Severe C19 presents a unique opportunity to probe the components of metabolic syndrome, especially those associated with the renin-aldosterone-angiotension system (RAAS). The ACE2 receptor ^2,3^ is the point of entry for SARS-COV-2^4^ and therefore was an early focus regarding RAAS hypertension therapy risks and C19 severity. ^5,6^ The question remains how, exactly, does the RAAS system impact C19. The role RAAS plays in hypertension (HT) makes RAAS an important target of HT drugs.^7–11^ Differences in HT severity^12^ and responsiveness to treatment modalities^12–16^ are associated with race and represented in current standard Clinical Practice Guidelines.^16^ How C19 affects RAAS among African Americans may offer more insight into how hypertension is expressed in this population.

Hyperlipidemia (HL), another component of metabolic syndrome, has been enigmatic in its impact for severe C19, with positive, neutral, and negative associations reported. Ryan et al^17^ reported a negative correlation with severity and claim to be the first to report a potential protective effect of statin use on COVID severity, though this effect may be entangled with race. Wu et al^18^ reports neutral risk for mortality, though they excluded controlled hyperlipidemic subjects. Kristen et al^19^ considered factors leading either to admission or mortality among diabetics, though severity as defined here was not reported. HL was found protective against mortality, and it suggested statins and fibrates as a possible reducer of lipids-induced inflammation. Atmosudigdo et al^20^ reported increased risk of severe C19 with HL in a meta-study.

In this study, we draw upon the Explorys^21^ dataset using both standard and novel^22,23^ epidemiological analysis techniques to explore the physiology of these risk factors elucidated by C19 associations, particularly through interactions with RAAS-targeted HT drugs,^7^ and the interaction of HL with other risk factors for severe C19. To address this epidemiological question, we developed an approach for Redescription-based Topological Data Analysis (RTDA) of clinical records to test our specific hypotheses and that can be generalized to address other epidemiological questions. Redescription analysis infers logical relationships among data features, while TDA examines the topology of these logical relationships yielding insights into possible multigenic pathways. While RTDA reveals possible multigenic pathways, we applied a method which performs cumulant-based network analysis (CuNA) to tie these features into sub-clusters or communities and obtain relative importance of each feature with measures of statistical significance. To this end, we found meaningful interactions between RAAS and severe C19.

## Materials and Methods

### Study Design

Explorys^21^ provides analytic tools and data, including de-identified electronic health records from emergency and clinical health services, and insurance records, compliant with the Health Insurance Portability and Accountability Act (HIPAA) and the Health Information Technology for Economic and Clinical Health Act (HITECH). The data are accessed from a guest portal with strictly controlled access, with a download prohibition of any primary de-identified records from the system. The portal hosts SQL access; our data snapshot was on 03/24/2021, then containing 55,972,084 records.

Explorys adapted the CDC NCHS coded racial and ethnic categories,^24^ which identifies Hispanic ethnicity primarily by country, while the proportion that reported themselves as “Hispanic” is a small fraction of the general population and were generally too small to test differential drug prescription choices or diagnoses, while African Americans were simply described.

Two sets of extracted records were accessed on the following criteria. Starting with a randomly selected 1,000,000 samples, 997,140 were retained having passed missingness. Lastly, we selected all the C19 patients plus an equal number of subjects not diagnosed with C19, yielding 539,523 subjects, of which 269,536 were C19 patients. These splits improve statistical power where the logistic regressions (LR) predict cases. The size of the randomly drawn set was selected to be similar in size to the C19 selected sets. The counts for relevant categories for all three sample sets are listed in Figure 1.

**Figure 1.**
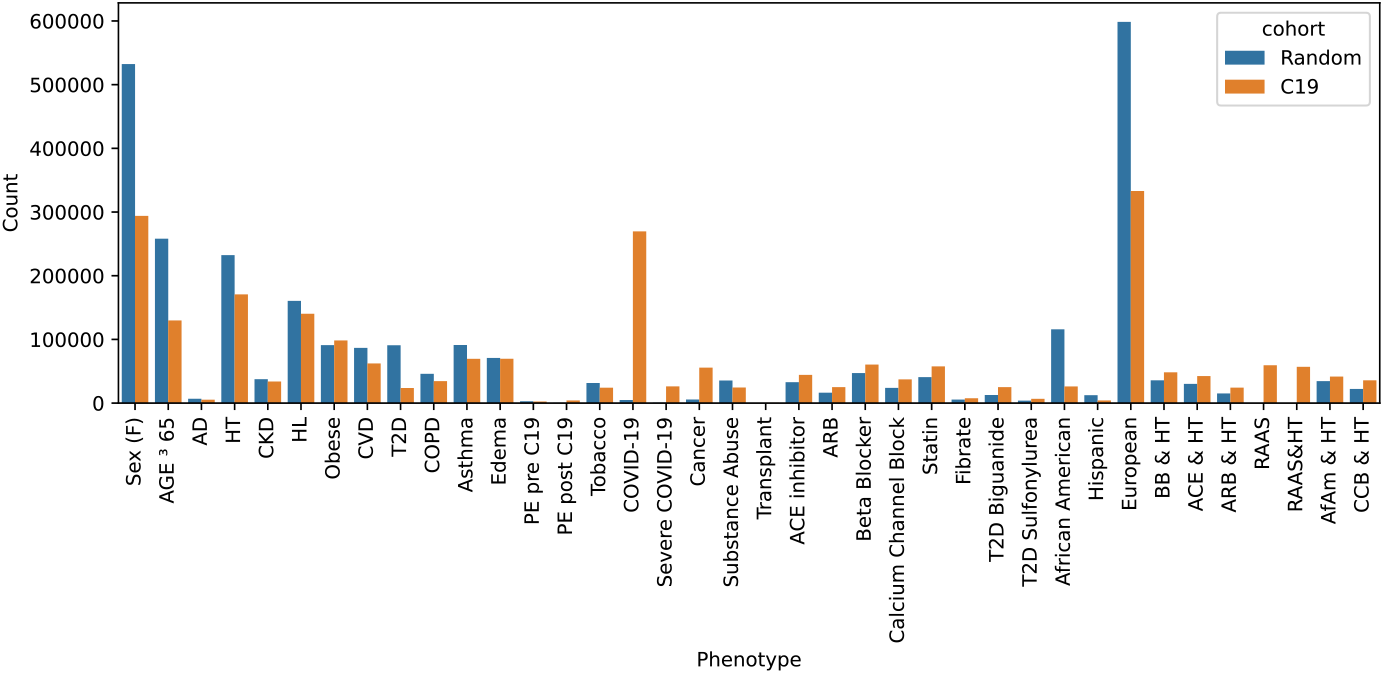
Two sample cohorts were drawn from the Explorys system: a random sample and all C19 patients plus an equal sized randomly selected non-C19 control group. The total counts indicate the number after dropping missing entries. Abbreviations include Alzheimer’s disease (AD), hypertension (HT), chronic kidney disease (CKD), hyperlipidemia (HL), cardiovascular disease (CVD), type 2 diabetes (T2D), pulmonary edema (PE), beta blocker (BB), calcium channel blocker (CCB), African Americans (AfAm).

### Redescription-based Topological Data Analysis (RTDA)

Joint cumulants^25^ show a number of useful properties. If the variables in the cumulant moments partition into blocks that are independent of each other, the joint cumulant vanishes,^26^ so contributions to joint cumulants that derive entirely from products of independent lower order cumulants cancel. In this sense, joint cumulants uniquely factor out contributions from lower order moments measuring purely n-way interactions. Significance was tested using Knuth’s “Algorithm P (shuffling).”^27,28^ permutation tests.^29^ In this approach, a null hypothesis is represented by randomly shuffled phenotype columns, from which a distribution of cumulants representing randomized associations are computed. The actual cumulants are compared with the distribution of random cumulant values.

Underlying biological processes and pathways are often distinguished by phenotypic combinations, from which inferences may be constructed. For example, Type II diabetes (T2D) patients have a high risk for HT. So T2D patients are nearly perfect subsets of HT patients. This may be rephrased as saying that T2D patients are captured by the same set as T2D intersected with HT patients. So logical relationships among phenotype variables may be represented in terms of equalities among intersections (logical “and”s) of these sets. These equalities are called “Redescriptions.” While allowing for misclassification, misidentifying stages of disease progression, and other errors, approximate equality of sets may be described in terms of Jaccard distances. In the case of subsets, the Jaccard distance yields the chances that a member in the putative subset is not contained in the other set. So, clustering the joint cumulants marking these clusters according to their Jaccard distances yields logical relationships between the phenotypes that may indicate underlying biological processes. These approximately equal sets are called “Fuzzy Redescriptions.”^30^ This analysis relies heavily on this formulation of redescriptions.

Complex diseases have multiple pathways yielding disease. If several collections of joint phenotype redescription clusters connect distinct phenotype conditions to disease along multiple pathways, it is evidence that multiple processes may be active leading to a pathology. We seek to recognize this situation by applying computation homology^31,32^ to identify topological loops among the pathways. The collections of deformed simplices represent connected redescriptions. Multiple paths connecting redescriptions will have holes in the cluster reflecting more than one pathway to disease. Thus, organizing the redescriptions into these homology groups is potentially informative of complex disease processes. We applied persistent homology analysis to explore homology groups as a function of connectivity thresholds, and their representative cycles used to characterize which redescriptions are relevant as markers along pathways identified as homological cycles.

### Cumulant-based Network Analysis (CuNA)

As a meta-analysis of joint cumulants we applied CuNA^33^ to extract higher-order relationships between the RAAS drugs, disease indications, age, sex, and COVID susceptibility and severity, respectively. CuNA counts higher-order relationships between pairs of conditions the set of clinical features from each of the cumulants, and embeds them into an easily digestible network where each node is a clinical feature and an edge between a pair corresponds to the number of times the pair were grouped together across the redescription clusters^22,33^. Thereafter, it performs community detection on statistically significant edges, where it obtains clusters from the nodes in the network and groups them together in meaningful functionally relevant groups. This process elucidates important modules of features which are interrelated, as well as the bridge nodes or communities which when removed results in connected components further highlighting their key role in understanding the shared etiology of the complex diseases related to RAAS. They also mark themes that emerge in redescription cycles.

## RESULTS

We applied standard LR and novel redescription-based topological data analysis (RTDA) epidemiological methods on patient cohorts to identify evidence of multiple pathways leading to pathology (complex disease) or other predictive and logical relationships among variates in clinical electronic records data extracted from Explorys. Two sample sets were extracted (Figure 1): *Random Cohort* (*n* = 997,140); *C19 Cohort* (case *n* = 269,536; control *n* = 269,987); after missingness exclusions. We found that severe C19 was a very small subset of the random cohort.

### Identifying relationships with logistic regression

LR was first used to investigate the impact of RAAS on the association between HT and severe C19, essentially probing RAAS by using severe C19 (Figure 2). Our LR findings also establish a familiar baseline for RTDA, which can be used to validate relationships captured by RTDA and highlight novel phenotypes LR is unable to discover. We probed metabolic syndrome associated drugs including RAAS and non-RAAS drugs for HT, biguanides and sulfonylureas for T2D, and fibrates and statins for HL. While we found HT is strongly associated with severe C19, we were able to isolate the contribution of the RAAS hypertension pathway to severe C19 by testing the interaction of HT with RAAS drugs (ACE inhibitors and ARBs), compared to non-RAAS drugs (beta blockers and calcium channel), on the association of HT with severe C19’s.

**Figure 2.**
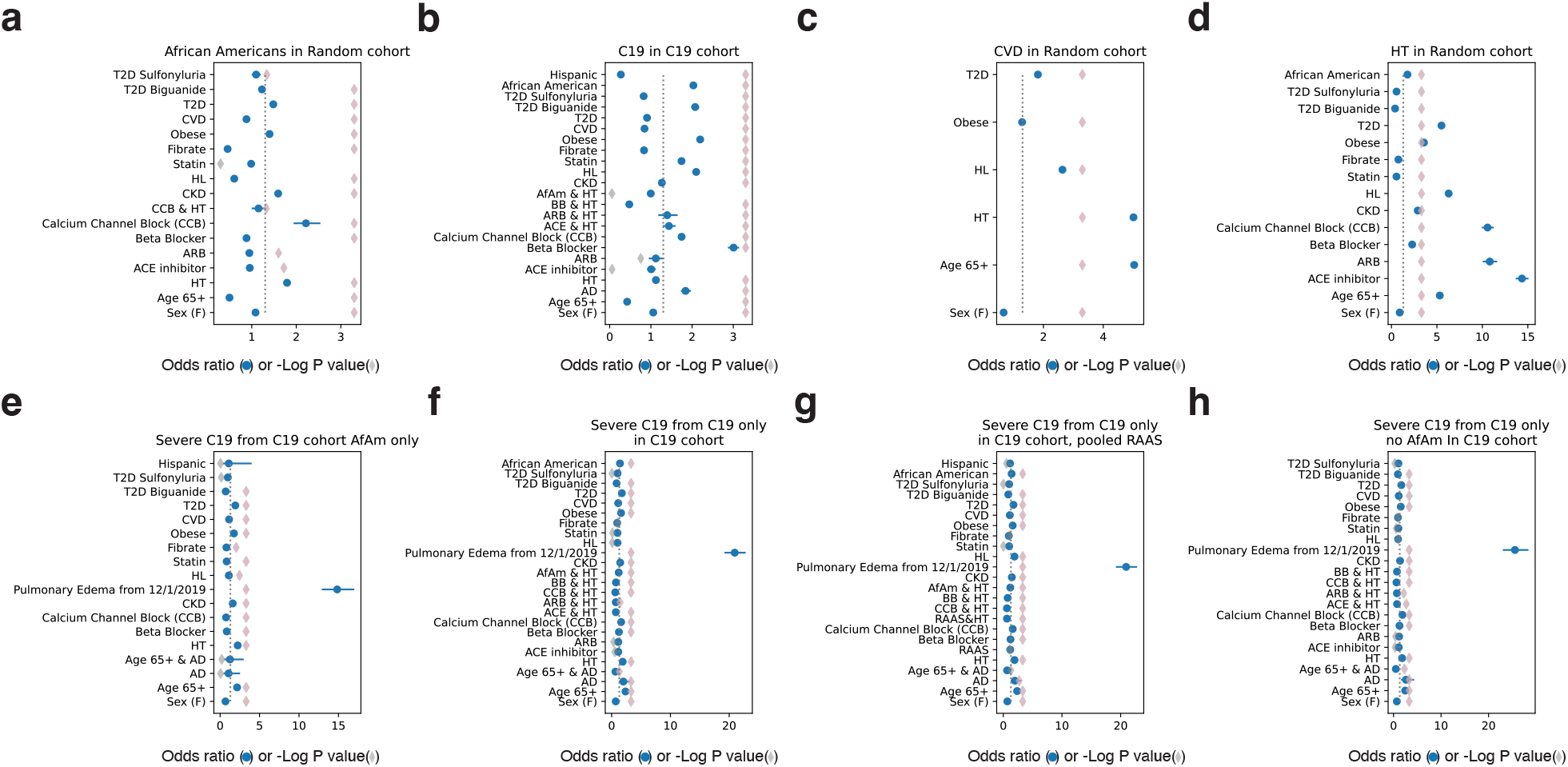
Stratified logistic regressions. a) Predicting African Americans (AfAm) in Random cohort. b) Predicting C19 in C19 cohort. c) Predicting CVD in Random cohort. d) Predicting hypertension (HT) in Random cohort. e) Predicting severe C19 in C19 cohort among AfAm. f) Predicting severe C19 from only C19 patients in C19 cohort. g) Predicting severe C19 from only C19 patients treated with RAAS drugs in C19 cohort. h) Predicting severe C19 from only C19 patients excluding AfAm in C19 cohort. Blue circles indicate the odds ratio. Blue bars are the 95% confidence interval. Diamonds are the –log(p value) and highlighted pink if p < 0.05 (gray dashed line).

Additionally, we observed distinct metabolic syndrome features among African Americans, including being prone to HT, T2D, and subsequent kidney failure (Figure 2a). Further, high lipid levels and prescribed treatments, as well as diagnosed cardiovascular disease, are underrepresented among African Americans in the Explorys set. Clinical practice guidelines for HT treatment for African Americans calls for calcium channel blockers (CCBs) plus another class of drug as first line therapy. Among African Americans in this dataset, CCBs are the dominant HT treatment, and RAAS drugs are poorly represented in those with severe C19. This result is not powered to resolve interactions between RAAS and hypertension, thus limiting resolution of RAAS drug influence among African Americans.

Logistic regressions comparing the impact of RAAS and non-RAAS drugs interacting with HT in predicting severe C19 in the C19 cohort were different (Figure 2f). The odds ratios for ACE inhibitors and ARBs were OR = 1.497 (95%CI: 1.09-2.05, p = 0.012) and 1.492 (0.93-2.39, p = 0.0958), respectively, and for beta blockers and CCBs 1.702 (95%CI: 1.47-1.96, p < 5 × 10^−4^) and 1.872 (95%CI: 1.37-2.55, p < 5 × 10^−4^). Pooled RAAS drug recipients, RAAS drug use on hypertensive association with severe C19, was OR = 1.405 (95%CI: 1.07-1.84, p = 0.0144). The beta blockers and CCBs for that regression showed 1.738 (95%CI: 1.51-2.00, p < 5 × 10^−4^) and 1.897 (95%CI: 1.39-2.59, p < 5 × 10^−4^), respectively. The odds ratio of hypertension by itself was 1.880 (95%CI: 1.80-1.97, p < 5 × 10^−4^). This suggests that treatment of hypertension with RAAS drugs may decrease risk of severe C19 compared to non-RAAS drugs.

### Redescription-based Topological Data Analysis (RTDA)

To identify evidence of multiple pathways leading to complex disease or other predictive and logical relationships among variates in clinical records data from Explorys, we performed RTDA. First, inferential information was collected, and we found 1940 possible compound predicates. Joint cumulants were used to identify n = 1825, P < 0.05 significant compound predicates. Each compound predicate is composed of subsets of subjects. Jaccard distances identify which subsets capture substantially the same subjects. For example, if subsets for descriptors *A* and *B* show *A* is close to the intersection of *A* and *B*, it follows that *A* is an approximate subset of *B*. In this case, if a subject satisfies predicate *A* then they are very likely to satisfy predicate *B*. Therefore, these clusters of predicates that capture the same subjects generate inferences. Such clusters of joint predicates are called “redescriptions.” Figure 3a presents a heatmap of Jaccard distances between joint predicates.

**Figure 3.**
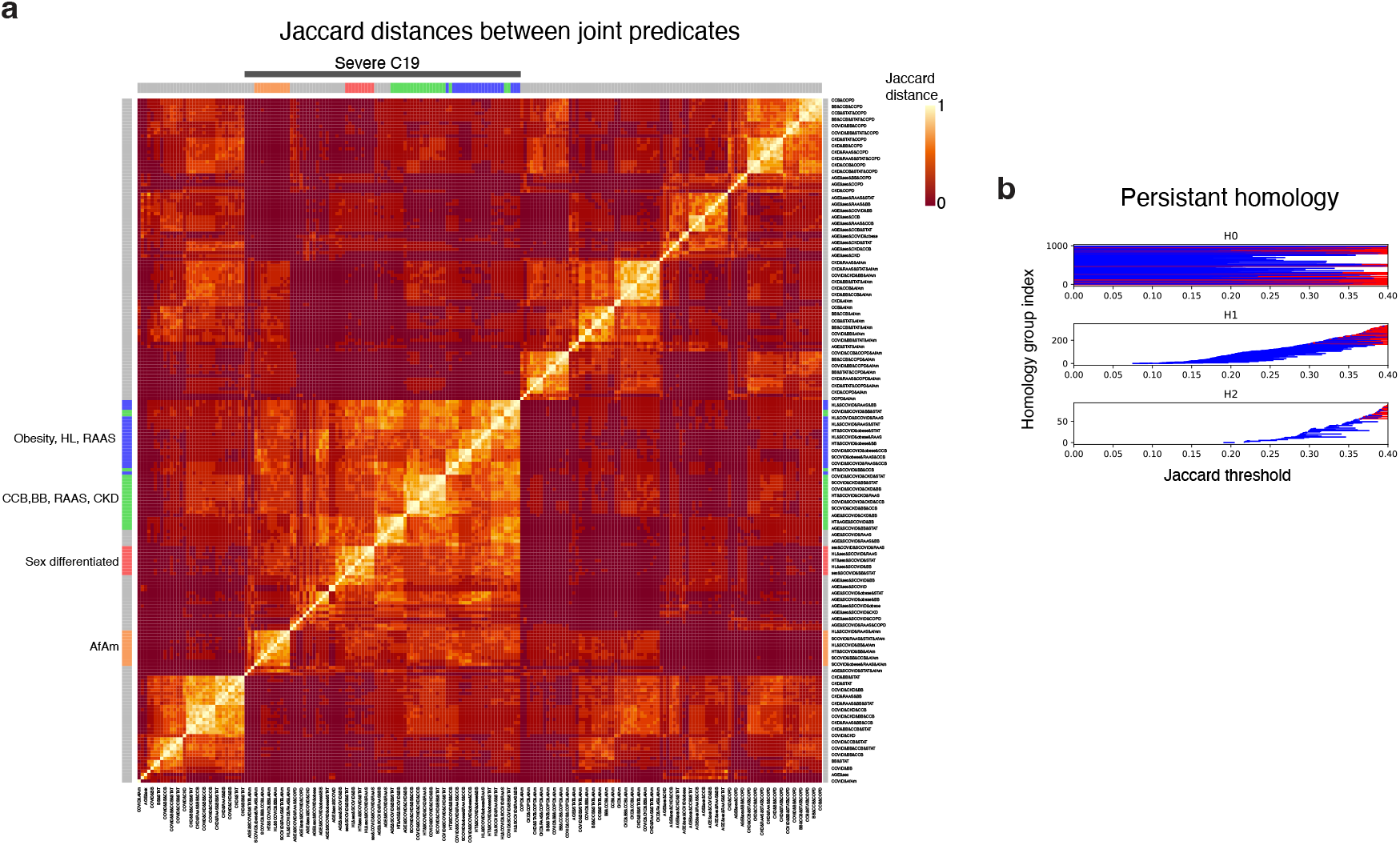
a) Jaccard distances between joint predicates. Severe C19 cluster is indicated with a gray bar (top). The primary homologous subclusters are indicated in blue, green, red, and orange. b) Barcode plots for the Jaccard distances shown in panel a. Abscissa is Jaccard threshold. Ordinate is homology group index ranked by Jaccard filtration threshold. The groups H0, H1, and H2 refer to dimensions 0, 1, and 2.

Computational homology was applied to compute persistence based on the Jaccard distances. For continuous variables, binary values were generated either from being above mean, or above clinically normal values as indicated in the Explorys records. The relationships among representative cluster connections are shown in Figure 3b. The horizontal bars mark connected diagrams (cycles, surfaces, volumes, etc., represented as ‘simplices’’). As the Jaccard filtration threshold is increased, new edges are added. The new edges connect previously distinct diagrams. Those distinct diagrams terminate at that filtration level, and a new diagram is created, often at a higher dimension, moving from *H*_0_ to *H*_1_ or from *H*_1_ to *H*_2_. With an upper bound of 0.5 applied to the Jaccard distance filtration, there were 998 cycles generated in *H*_1_. The four leading cycles capturing severe C19 segregates strongly in the heatmap, and largely include most of the severe C19 cases marked by the colored sidebars (Figure 3a).

Figure 4 displays some of the largest representative cycles within the homology groups identified by the persistent homology algorithms highlighted among the plotted Jaccard distances (Figure 3a). The cycles were selected to highlight inclusions involving severe C19 and three HT drugs, including ACE inhibitors, beta blockers, and ARBs, while the fourth figure involves African Americans. The largest Jaccard distance within a cycle defines the birth of the homology group bars (Figure 3b). The largest distances in each cycle were: 0.254 – ACE inhibitor cycle (Fig. 3a, 4a, red); 0.370 – age, chronic kidney disease (CKD), CCB, hyperlipidemia/statin cycle (Fig. 3a, 4b, green); 0.223 – CCB, obesity, and hyperlipidemia cycle (Fig. 3a, 4c, blue); 0.321 – African American cycle (Fig. 3a, 4d, orange).

**Figure 4.**
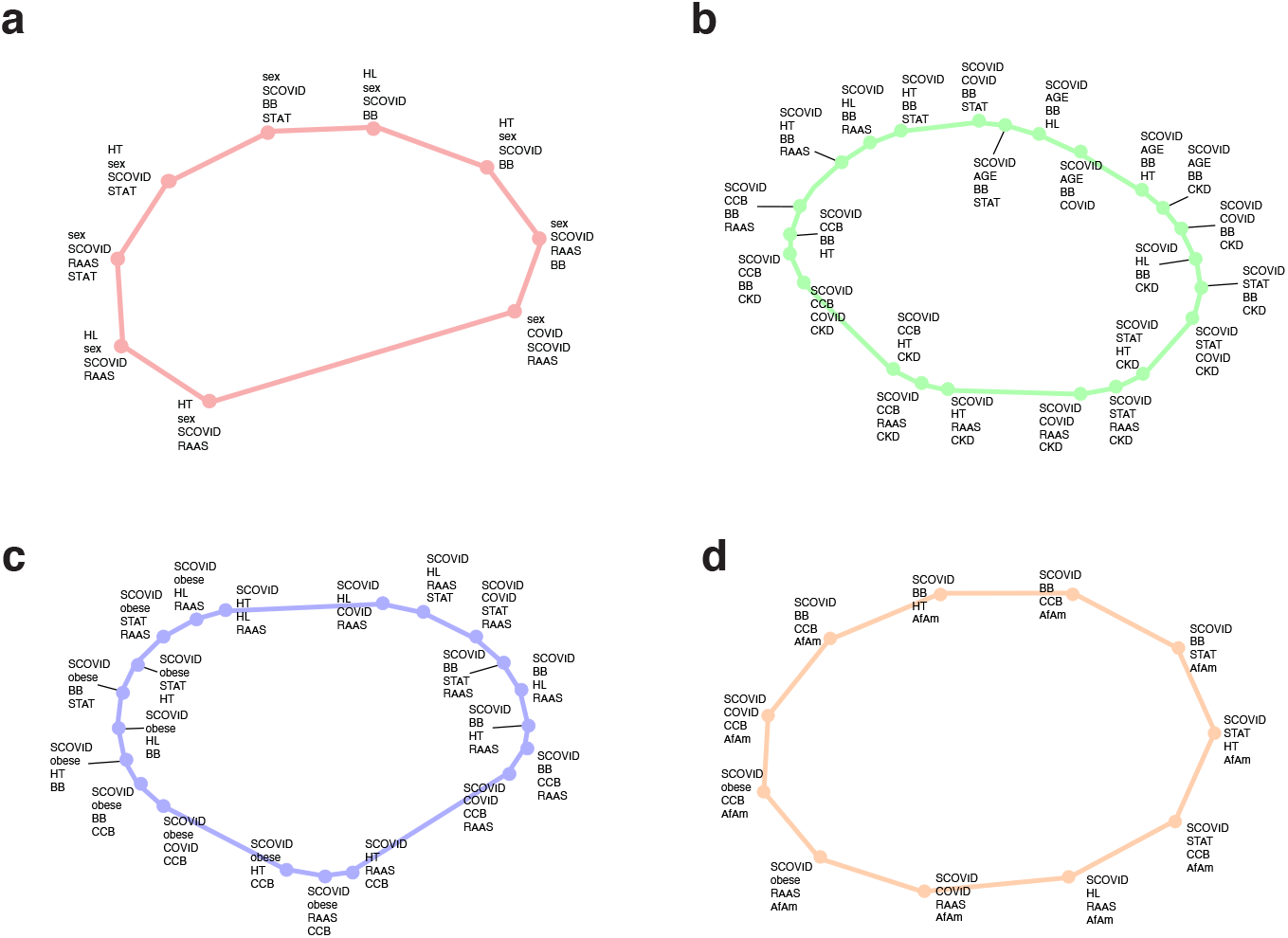
Representative cycles corresponding to side-bar color codes in Figure 3a selected to highlight inclusions involving severe C19 and three hypertension drugs (ACE inhibitors, beta blockers, and ARBs), as well as African Americans. a) ACE inhibitor cycle. b) CCB, Age, and CKD cycle. c) CCB, obesity, and HL cycle. d) African American cycle

The redescription clusters tend to sit in segments connected on the larger homological cycles, suggesting multiple pathways associating these groups with each other. These clusters are:

- Red: sex differentiated responses.
- Green: CCB, BB, and RAAS and patterns not selected by RAAS but heavily identified by other hypertensive drugs, connected to patients with chronic kidney failure with RAAS, elderly patients with RAAS, and a cluster with statin treatments.
- Blue: resembles green cycle, but with more focus on obesity and HL. It shows several sets of redescriptions connected into a multi-pathway cycle. A connecting feature of all these redescriptions is the lipids cycle – either obesity, statin prescriptions, or hyperlipidemia.
- Orange: African Americans with severe C19 with CCB prescription, hyperlipidemia, and BB prescription. Thus, RAAS components stand out as tending to be associated with obesity, hyperlipidemia, and statin treatment in severity of C19 in African Americans.

Each of these cycles implicate HL as a factor among severe C19 patients, even though LR did not identify a first order association between HL and C19 severity. Our LR predicting severe C19 from C19 patients only in the C19 cohort while pooling ARB and ACE-inhibitor RAAS drugs indicated an OR=1.005 (95%CI: 0.97-1.04, p= 0.793) (Figure 2g), consistent with other reports that hyperlipidemia is unimportant to severe C19^17–20^. To understand the signals RTDA detected, we performed LRs stratified by hyperlipidemia (Figure 5). We found that hyperlipidemia significantly reduced the impact of other metabolic syndrome conditions on the severity of C19, including being male, age 65 or over, hypertension, chronic kidney disease, pulmonary edema during the year of the C19 infection, obesity, and type-II diabetes. This protective effect is in keeping with some the published work on HL with C19^17,19^. The only aggravated interaction was with the impact of cardiovascular disease on C19 severity (Figure 5).

**Figure 5.**
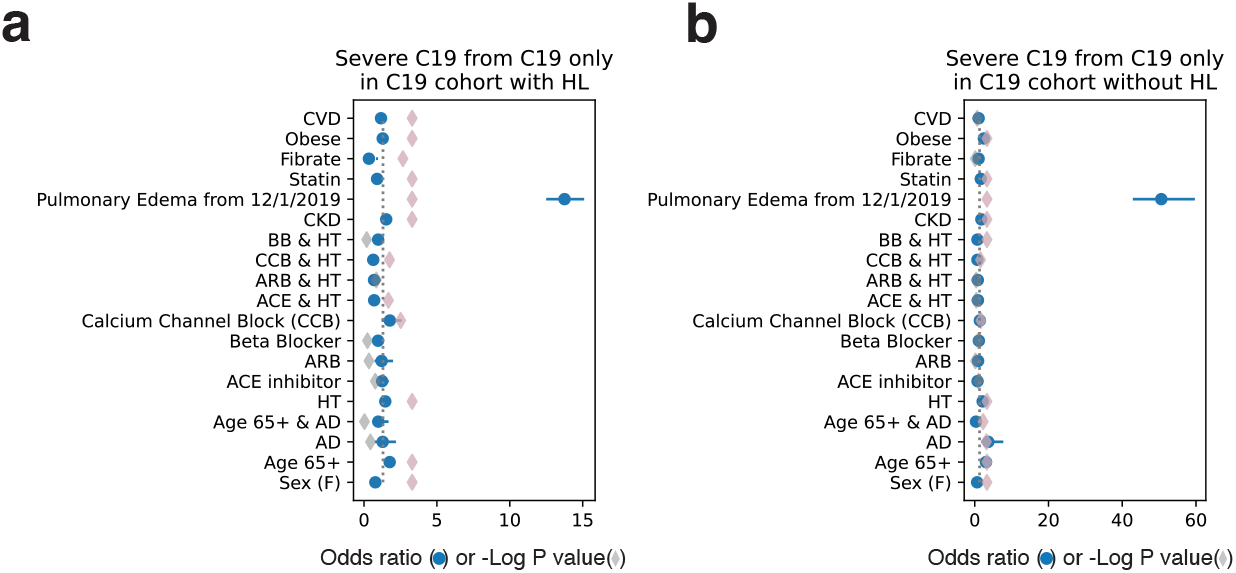
Stratified logistic regression predicting severe C19 computed on C19 only patients from the C19 cohort with (a) and without (b) hyperlipidemia. Blue circles indicate the odds ratio. Blue bars are the 95% confidence interval. Diamonds are the –log(p value) and highlighted pink if p < 0.05 (gray dashed line).

### Networks and Communities

Using CuNA, we derived a network that highlights the importance of pairwise associations across multiple sets of interactions represented within the individual joint cumulant clusters. For example, while HL was not itself significant to severity of C19, it interacted with other factors to enhance their impact on severity. CuNA identifies those contributions that a simple LR might miss.

To test the significance of the CuNA network (Figure 6), we evaluated the interactions over a range of p-values *p* = (0.01, 10^−12^}, incrementing it by 10^−1^ and z-scores *z* = (2, 4) incrementing by 0.5. We generated 50 different networks (for varying values of *p* and *z*) and retained edges that occurred in all networks (Figure 6). The width of the edges corresponds to the number of times a pair of nodes appeared together in the cumulant groups and reflects their pairwise affinity. As a result, we conclude that these interactions are robust across various thresholds of significance. Further network analysis highlighted RAAS as one of the top influential nodes (Table 1) in the network obtained by CuNA (Figure 6). The importance of each node is calculated as an aggregate score defined by the mean of the ranks of different network centrality measures: betweenness centrality (number of unweighted shortest paths between all pairs of nodes in the graph that passes through each node); eigenvector centrality (relative importance of a node as compared to its neighbors); degree (number of edges connected to the node); Voterank (a ranking based on a voting scheme between the neighbors of each node); and information centrality, or current flow closeness, (based on effective resistance between nodes in a network).

**Table 1.**
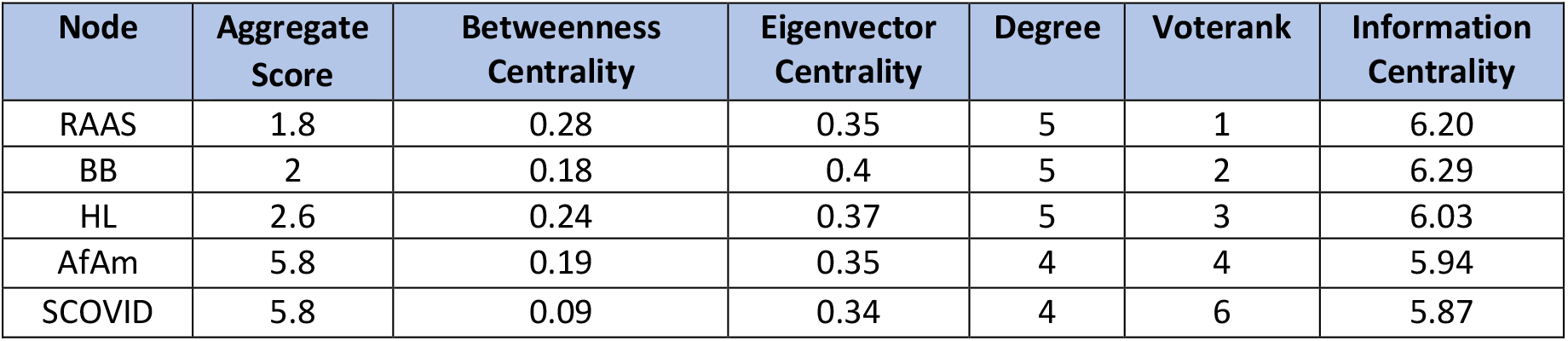
Top 5 nodes ranked by an aggregate score of different network centrality measures.

**Figure 6.**
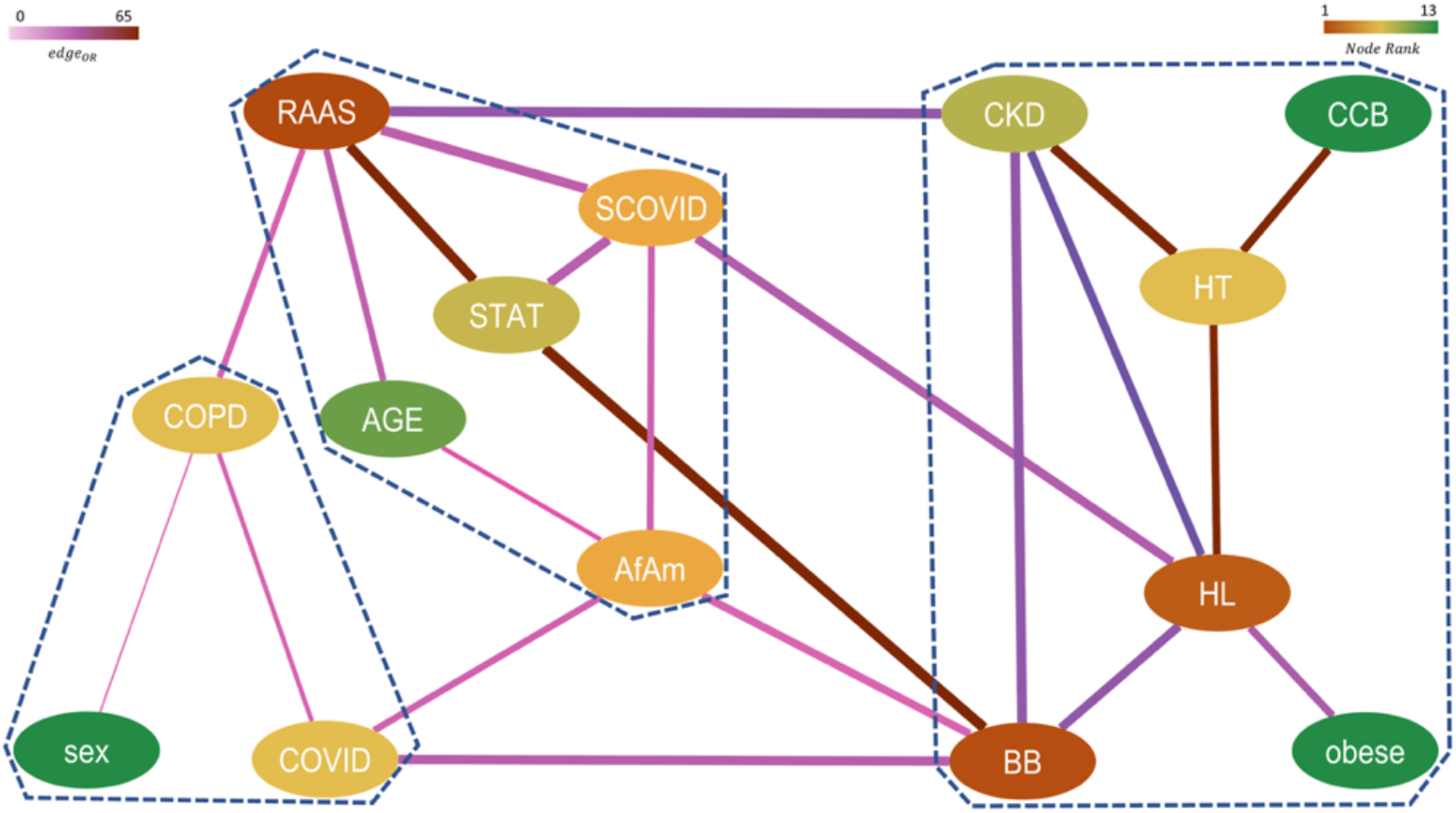
Network representing higher-order significant interactions between different features with nodes colored by their relative rank (gradient of brown to green corresponds to higher to lower rank) and edges colored by their respective pairwise ORs (gradient of light to dark corresponds to low to high OR) and edge width indicates the strength of the connection between them. The communities obtained from this network are marked by dashed lines.

We observed three communities from the significant cumulant groups using a greedy modularity-based algorithm^34^ (modularity score = 0.78). The final communities are marked by a dashed line in the CuNA network (Figure 6).

- RAAS, Severe Covid-19 (SCOVID), African Americans (AfAm), Statins (STAT) and age.
- Beta-blockers (BB), CKD, obesity (obese), HL, HT and CCB.
- Susceptibility to C19 (COVID), Chronic Obstructive Pulmonary Disorder (COPD) and sex.

We observe the highest ORs are between the pairs {HT, CCB}, {HT, CKD}, {RAAS, STAT}, and {BB, STAT}. The CuNA analysis is supported by the recovery of well-known clinical relationships. For instance, CCB and HT have the highest OR in the network. The community of BB, HL, and obesity follows from the fact that beta-blockers can have an adverse effect on blood lipids^35–37^ and obesity is a well-documented side effect of beta-blockers.

## Discussion

Clinical electronic medical records data can provide a sampling of how complex disease conditions, such as metabolic syndrome, may interact with emergent syndromes, such as C19. Utilizing RTDA and CUNA, we discovered higher-order combinatorial relationships between the cluster of conditions associated with metabolic syndrome and severe C19. Such methodologies also probed how C19 severity interacts with RAAS pathways by considering drugs and susceptibility to severe C19. We observed a protective effect for severe C19 by RAAS-associated HT therapy and for HL with other severe C19 risk factors, as well as detected the impact of race on susceptibility.

Differential impact of hypertension on severe C19 between RAAS vs. non-RAAS targeting medication users suggests that HT pathways involving RAAS are distinct in how they participate in the induction of severe C19. While RAAS drug prescription was a risk for severity in itself among C19 patients registered in Explorys, the risk for severe C19 due to hypertension is reduced for patients under RAAS therapy compared to CCBs and beta blockers. This protective effect was also shown in Bezabhe et. al.^11^, though they report no overall direct impact of RAAS inhibitors distinct from the comorbidities. In our study we show there is a distinct interaction of RAAS drugs with the impact of hypertension on severe C19.

RTDA, recovered the severe C19 patients as a distinct cohort, predominantly characterized by cycles involving metabolic syndrome. Most of the patients show involvement of multiple components of metabolic syndrome treated with multiple drug therapies. Moreover, hyperlipidemia is implicated in severe C19. This finding highlights the involvement of higher order interactions since higher order cumulants involving hyperlipidemia would have cancelled if the associations could have been accounted for by lower order cumulants. The observed significance for these cumulants, verified by logistic regression, indicates that hyperlipidemia impacts the interaction of hypertension and other metabolic syndrome components with C19 severity. The direct association between hyperlipidemia with C19 severity was weak. The finding by RTDA of HL’s connection to severe C19 led us to revisit the role hyperlipidemia played that had been missed by a first order LR. An analysis of other risk factors stratified by hyperlipidemia showed shifts in the impact of those risk factors on C19 severity (Figure 5). This is further corroborated by the CuNA analysis showing hyperlipidemia, BB, CKD, obesity, CCB, and HT appearing in the same community, primarily focusing on comorbid diseases such as HT, and CKD and hypertensive drugs such as BB and CCB, all of which are strongly related to hyperlipidemia^35–38^.

While the relationship between HL and HT is expected for the general population, RTDA reveals this to be an important consideration among African Americans, where the condition tends to show lower diagnosis rates and treatments in the Explorys dataset. RTDA identifies these factors in multiple component cumulant clusters involving African Americans. While LR indicated CCBs are a treatment of choice for African Americans, and picked up other distinctive characteristics, such as lower diagnosed rate of HL, statin or fibrate prescriptions, or diagnosis of CVD, it does not adequately characterize the interconnected relationship between these factors. Whereas in the RTDA cycles all components of the cycle are significant and relevant to severe C19. Furthermore, African Americans appeared among the top five most significant nodes in the CuNA network highlighting the importance to study this population in relation with C19 severity and metabolic syndromes (Table 1) and it belonged to a community with elevated severity of C19. Lastly, the topological cycles and redescriptions suggested several novel relationships that could be tested by, and quantified using, LR to assess their validity and measure the association strengths.

## Conclusions

RAAS plays a central role in the CuNA network (Figure 6) being the most influential node (Table 1). The specific community containing RAAS also includes statins and severe C19, which is meaningful as there have been conflicting reports as to the role of HL with severe C19. This community, which also includes age and African Americans, emphasizes the previously known interconnection between severe C19 with African Americans and the elderly. When applying methods such as RTDA to clinical health records data, it is important to keep in mind that the results will reflect clinical practice guidelines and any potential underlying racial disparities, such as the relationship between severe C19 and African Americans.

RTDA enabled the discovery of higher-order relationships beyond typical first order logistic regression models exploring epidemiological questions. Using RTDA, we sought to understand how C19 can be leveraged to understand the relationship between the risk factors for C19 and the RAAS component of metabolic syndrome, and note that severe C19 may be used as a probe into RAAS pathways in metabolic syndrome. We found that the RAAS complex was important due to involvement of chronic kidney disease, and the trend towards suppression of the association of hypertension with severe C19 by RAAS drugs vs non-RAAS drugs, though those associations are not statistically significant. Redescriptions and homological cycles did not tend to pick up such associations. Instead, metabolic syndrome and related conditions appear to be important to severe C19, but with some variations depending on comorbidities or distinctive lineages. Specifically, some variations show up within African Americans compared to other groups, which is also reflected in differences in clinical practice guidelines. This study on the severity of C19 highlights the potential for topological data analysis to aid in epidemiological studies looking at specific relationships to uncover novel connections beyond the power of traditional analyses.

## Data Availability

All data produced in the present study are available upon reasonable request to the authors.

